# Standardization of CFSE and Ki-67 based lymphocyte proliferation assays in the absence of thymidine-based methods among children with nephrotic syndrome in south India: A pilot study

**DOI:** 10.1101/2025.08.12.25333468

**Authors:** Georgie Mathew, Poornima Saravanan, Chanduni Syed, Akilandeswari Eswaran, Ajith Kumar, Esra Oliver, Sathish Kumar L, Leenu Lizbeth Joseph, Indira Agarwal, Ramya Madhavan

## Abstract

**Background:** Lymphocyte proliferation assays are essential for assessing cellular immune responses, especially in immunocompromised individuals. Traditional thymidine-based methods have limitations such as radioactive hazard and limited accessibility in low-resource settings. This study aimed to optimize and validate carboxyfluorescein succinimidyl ester (CFSE) and Ki-67 based non-radioactive proliferation assays for potential application in children with nephrotic syndrome receiving corticosteroids.

**Methods:** Peripheral blood mononuclear cells (PBMCs) from a healthy adult donor were used to optimize assay parameters including CFSE concentration, cell density, and stimulation kinetics. PBMCs from 20 pediatric participants were then stimulated with phytohemagglutinin (PHA). CFSE-labelled cells were analysed on day 6 for cell division, while Ki-67 expression was measured at 72–75 hours post-stimulation by flow cytometry. Proliferation was quantified as the percentage of proliferating cells after subtracting background from unstimulated controls.

**Results:** The optimal CFSE concentration was 2.5□μM at a cell density of 1□×□10□ cells/mL. Ki-67 expression was stable between 69–96 hours, enabling processing flexibility. A cut-off of >50% proliferation was used for both assays. Among the pediatric samples, a moderate but statistically significant correlation was observed between CFSE and Ki-67 expression assays (Spearman’s r = 0.57, p = 0.01), indicating concordance.

**Conclusion:** This pilot study demonstrates the feasibility and reproducibility of CFSE and Ki-67–based assays especially with lower PBMC counts from pediatric population. These methods offer reliable, non-radioactive alternatives for immune monitoring in nephrotic syndrome cohorts. Validation in larger and diverse populations is recommended before broader implementation.

## Introduction

Lymphocyte proliferation assays (LPA) are crucial for assessing the safety and efficacy of live vaccines in immunosuppressed individuals, as it helps determine the functionality of their immune system and their ability to mount an adequate response to the vaccine [1]. This assay measures the ability of lymphocytes to multiply in response to a specific antigen, indicating the individual’s cellular immunity [2]. Traditionally, LPAs are performed with tritiated thymidine incorporation, and require radioactive handling, high costs, over-analyses and high cell counts, making them particularly unsuited for pediatric populations [2,3].

Several alternative methods for assessing lymphocyte proliferation have been developed and are widely used in clinical and research settings. Among these, colorimetric assays such as the MTT-based assay and dye dilution techniques using carboxyfluorescein succinimidyl ester (CFSE) or CellTrace™ Violet (CTV) are commonly employed [4,5]. These fluorescent dyes are incorporated into cells and progressively diluted with each cell division, generating distinct daughter peaks detectable by flow cytometry. More sensitive techniques like 5-bromo-2’-deoxyuridine (BrdU) and 5-ethynyl-2’-deoxyuridine (EdU) incorporation assays allow direct labeling of newly synthesized DNA. However, one potential disadvantage of both BrdU and EdU is the damage to daughter cell and induction of mutations, making them less suitable for downstream or long-term functional assays [6,7]. Ki-67, a nuclear protein expressed during all active phases of the cell cycle (G1, S, G2, and M) but absent in resting (G0) cells, offers a non-radioactive, scalable, and relatively safer alternative for assessing cell proliferation, particularly in flow cytometry–based applications [8]. Ki-67 can detect proliferating lymphocytes with a sensitivity greater than BrdU incorporation and has a sensitivity similar to dye dilution methods such as Oregon green (OG) which is a CFSE derivative [8].

Thus, we selected CFSE and Ki-67 staining based mitogen stimulation assays to assess lymphocyte proliferation in this pilot study. We selected CFSE and Ki-67 based mitogen stimulation assays to assess lymphocyte proliferation in this pilot study, as these methods offer complementary insights. CFSE tracks cell division through dye dilution over time, while Ki-67 serves as an intracellular marker of active proliferation. Although these methods are increasingly used in the evaluation of immunodeficiencies, detailed and standardized protocols for CFSE and Ki-67–based proliferation assays in pediatric populations particularly from South Asia remain limited. This pilot study aims to optimize and validate these two complementary approaches using cryopreserved PBMCs from children in South India, including both healthy controls and children with nephrotic syndrome on corticosteroids. The findings will inform a future studies on evaluating the safety and immunogenicity of live attenuated vaccines in children with nephrotic syndrome receiving corticosteroid therapy.

## Materials and Methods

### Study Design and Sample Collection

This pilot study aimed to develop and validate lymphocyte proliferation assays using CFSE and Ki-67 in pediatric peripheral blood mononuclear cells (PBMCs) with and without nephrotic syndrome, in the absence of thymidine-based methods. A healthy adult volunteer provided a blood bag sample from which assays were performed for protocol optimization. The protocol optimization involved staining concentration and stimulation time for CFSE, time kinetics and volume titration, cell density for Ki-67. Subsequently, PBMCs were isolated from 18 children recruited from pediatric nephrology and hemato-oncology departments in a tertiary care institute from South India. Informed consent was obtained from guardians of all participating children, and the study was approved by the institutional ethics committee (IRB Min. No. 2411130).

Participants were classified into four groups based on clinical status: well children, children with nephrotic syndrome (currently being administered corticosteroid therapy or off) and 2 children immediately post-autologous stem cell transplant (for T cell acute lymphoblastic leukemia and metastatic neuroblastoma). Demographic data, treatment history, and relevant clinical information were recorded. PBMCs from all participants underwent both CFSE and Ki-67 proliferation assays under identical culture and staining conditions. Table 1 shows the participant characteristics.

**Table 1:** Clinical and laboratory characteristics of the participants at baseline. The values are represented as median with interquartile range.

| **Parameter** | **Total cohort**<br>**(n = 20)** | **Well children**<br>**(n = 3)** | **NS off steroids**<br>**(n = 3)** | **NS on steroids**<br>**(n =12)** | **Immunodeficient state**<br>**(n = 2)** |
| --- | --- | --- | --- | --- | --- |
| **Age (years)** | 8 (5.8, 11) | 6 (5, 9) | 11 (11,12) | 8 (5.8, 11) | 6, 5 |
| **Males (%)** | 13 (65) | 2 (67) | 1 (33) | 9 (75) | 1 (50) |
| **Hemoglobin, g/dL (n = 19)** | 13.3 (12.4, 13.9) | 14.1 (13.3, 14.1) | 14.1 (13.6, 14.5) | 13.3 (12.5, 13.7) | 10.3, 12.1 |
| **Total WBC counts/mm<sup>3</sup> (n = 14)** | 9350 (6150, 13225) | 8050 (7025, 9075) | 7350 (6625, 8075) | 12950 (9075, 14875) | 6900, 2800 |
| **Neutrophils, % (n = 14)** | 59.5 (51.8, 68.8) | 51 (47, 55) | 70 (67,73) | 59 (48.8, 68.2) | 78, 57 |
| **Absolute neutrophil count, /mm<sup>3</sup> (n = 14)** | 5481 (3469, 6626) | 4269.5 (3425, 5114) | 5232 (4504, 5960) | 6009 (4257, 8733) | 5382, 1596 |
| **Lymphocyte %, (n = 14)** | 27 (20, 34) | 35 (33, 37) | 19.5 (19.2, 19.8) | 29.5 (25.2, 37) | 4, 20 |
| **Absolute lymphocyte count, /mm<sup>3</sup> (n = 14)** | 2006 (1264, 3493) | 2736 (2538, 2933) | 1426 (1303, 1549) | 3475.5 (1613, 4197) | 276, 560 |
| **CD4+ T-cells counts (n = 2)**<br>**CD4+ T cell percentage (n = 2)** | 1635, 2174<br>41, 44 |  | 1635, 2174<br>41, 44 |  |  |

### PBMC Isolation

Venous blood was collected in heparinized blood collection tubes (BD vacutainer 455051) and processed within two hours of collection. Whole blood was spun at 200xg for 10 min to separate plasma from cells. PBMCs were isolated on a Ficoll-Paque density gradient medium (Sigma, Histopaque-1077, 10771-6×100ml), washed with 1X PBS (made from 10x PBS-Thermo Fisher, 70011044) and resuspended in 90% FBS (Thermofisher,10270106) and 10% DMSO (Sigma Aldrich, D2650). Cells were stored in liquid nitrogen in aliquots of 5-10 million per vial and thawed as needed for experiments.

### Optimization of CFSE Staining

CFSE (Thermo Fisher, C34554) staining was optimized using PBMCs from the adult volunteer. To identify the optimal CFSE concentration for lymphocyte proliferation assays, PBMCs were stained with CFSE at four concentrations: 1.25 μM, 2.5 μM, 5 μM, and 10 μM. Cells were resuspended at a concentration of 10 × 10□ cells/mL in PBS containing 1% FBS and incubated with the respective CFSE concentrations in 5 mL round-bottom polystyrene tubes. The staining was performed for 10 minutes at room temperature in the dark, with gentle mixing every 2 minutes to ensure uniform dye uptake. The reaction was quenched by adding 4 mL of chilled complete medium, followed by two washes with complete RPMI (cRPMI) to remove excess dye. Labelled cells were stimulated with phytohemagglutinin at 1% v/v (PHA-M, Thermo Fisher,10576015) and incubated at 37°C in the CTS OpTmizer T Cell Expansion SFM containing glutamine (Thermo Fisher, A1048501), 5% CO□ for 6 days. Flow cytometry was used to assess proliferation by evaluating CFSE dilution. After we finalised the CFSE concentration, we also evaluated the optimal duration of stimulation for effective assessment of CFSE-based lymphocyte proliferation by analyzing samples at 3, 5, and 6 days post-stimulation. To assess viability and ensure accurate gating of T cells, cells were stained with Fixable Viability Stain 700 (FVS700, BD Biosciences, 564997), followed by surface staining with anti-CD3 antibody (CD3-APC, BD Biosciences, 340440) (Supplementary figure 1). To assess repeatability, the assays were performed in duplicate on two separate days using the same sample (Supplementary Figure 2).

### Optimization of Ki-67 Staining and Time Point Determination

For Ki-67 assays, PBMCs were stimulated with PHA-M (at 1% v/v) and harvested at multiple time points (24 h, 69 h, 72 h, 75 h, 78 h, 81 h, 96 h, and 120 h) to determine the kinetics of Ki-67 to assess proliferation. After PHA stimulation, PBMCs were first stained for viability using Fixable Viability Stain 700 (FVS700, BD Biosciences, 564997) to exclude dead cells. This was followed by surface staining with anti-CD3 antibody (CD3-V500, BD Biosciences, 561416) to identify T cells. Cells were fixed and permeabilized using a commercial fixation/permeabilization kit (Cytofix /Cytoperm Soln Kit, BD Bioscience-554714), followed by incubation with anti-Ki-67 antibody (clone B56, 2.5 μL/test, BD Biosciences-567719). Ki-67 was also titrated at varying volumes starting from 5 μL, 2.5 μL, 1.25 μL, 0.625 μL and 0.3 μL to determine the optimum volume for staining.

### Cell Seeding Density Optimization

To determine the optimal cell density for CFSE and Ki-67-based lymphocyte proliferation assays, PBMCs were seeded at four concentrations: 5 × 10□, 2.5 × 10□, 1 × 10□, and 0.1 × 10□ cells/ml. Proliferative responses and staining quality were evaluated across these densities to identify a concentration that provided reliable assay performance without compromising cell viability. This optimization was necessary since traditional thymidine-based proliferation assays typically require minimum of 5 million cells per participant.

### Analysis

All stained samples were acquired using a BD FACS Symphony flow cytometer. A minimum of 0.2 million lymphocyte events were collected per sample. Data were analyzed using using FlowJo, LLC (version 10.8.1). CD3□ T cells were identified by gating on lymphocytes based on forward and side scatter properties, followed by exclusion of dead cells using viability dye and selection of CD3-positive events. Proliferation was assessed using two approaches: (i) CFSE dilution, visualized as multiple peaks corresponding to successive cell divisions, and quantified using the proliferation index generated by FlowJo; and (ii) percentage of Ki-67□ CD3□ T cells after subtraction from their matched unstimulated controls.

Proliferation data were summarized as medians and interquartile ranges. Correlation between Ki-67 and CFSE was assessed by Spearman correlation. A p-value <0.05 was considered statistically significant.

## Results

### Optimization of CFSE Staining

To identify the optimal CFSE concentration for proliferation tracking, 1□×□10□ PBMCs from the healthy adult was labelled with increasing concentrations of CFSE (1.25 μM, 2.5 μM, 5 μM, and 10 μM) and stimulated with PHA for 5 days. Flow cytometric analysis revealed that 2.5 μM CFSE provided the best compromise between fluorescence intensity and cell viability. At this concentration, distinct peaks representing successive cell divisions were observed following PHA stimulation. Higher concentrations, such as 5 μM and 10 μM, resulted in elevated background fluorescence and reduced cell viability. Therefore, 2.5 μM was selected for all the CFSE-based proliferation assays (Figure 1 A-E). Subsequently, we also determined the number of days required to assess the CFSE based cell proliferation and found that 5 days (6^th^ Day post stimulation) were required to get the maximum proliferation (Figure 2)

**Figure 1:**
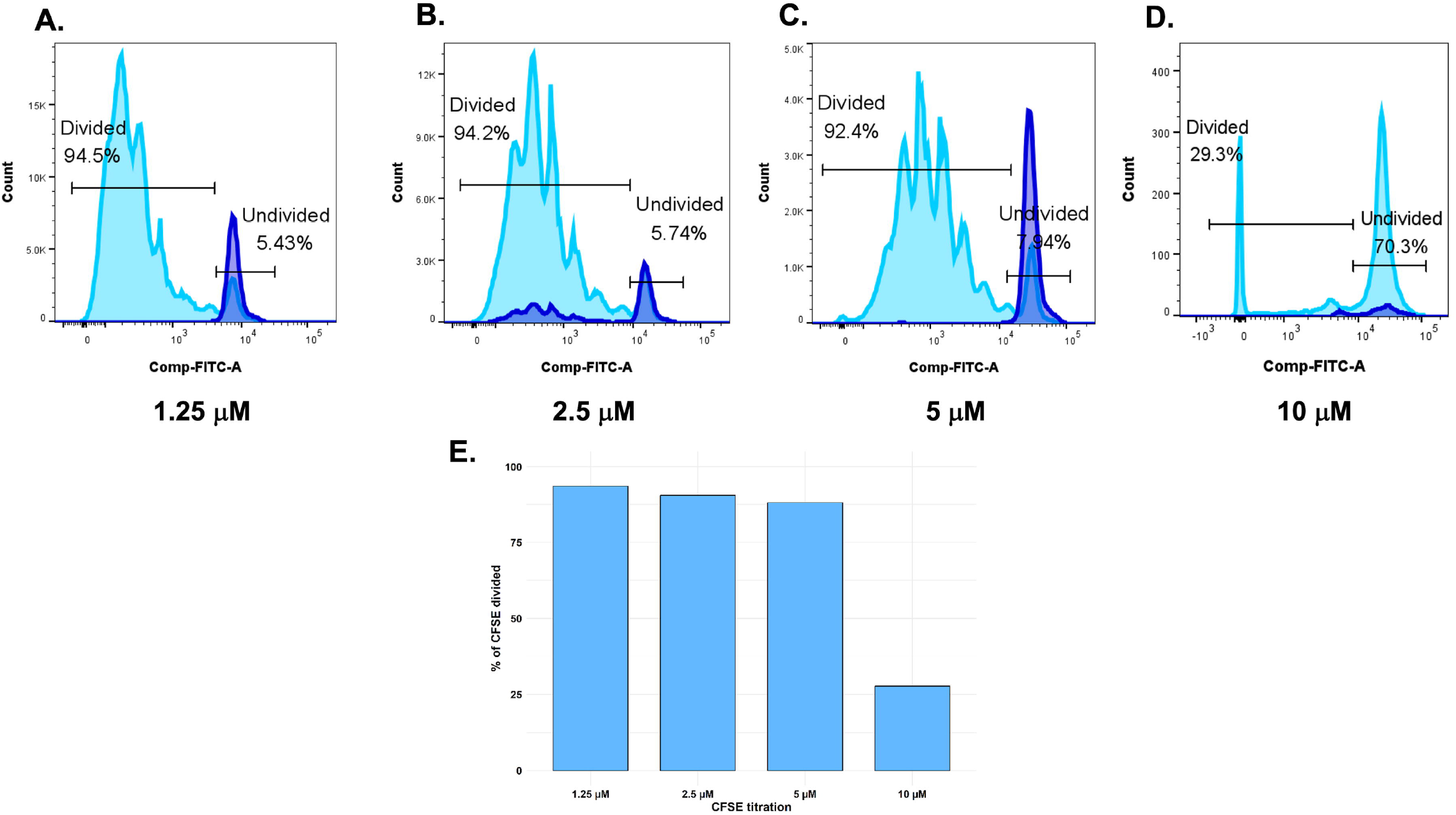
CFSE titration to determine optimal concentration for T cell proliferation assay. Panels A–D show histograms of CFSE-stained T cells after 5 days of stimulation with PHA, using different CFSE concentrations: (A) 1.25□μM, (B) 2.5□μM, (C) 5□μM, and (D) 10□μM. CFSE fluorescence intensity decreases with each cell division, enabling visualization of proliferating cells. (E) Bar graph depicting the percentage of CFSE low (divided) T cells corresponding to each CFSE concentration.

**Figure 2:**
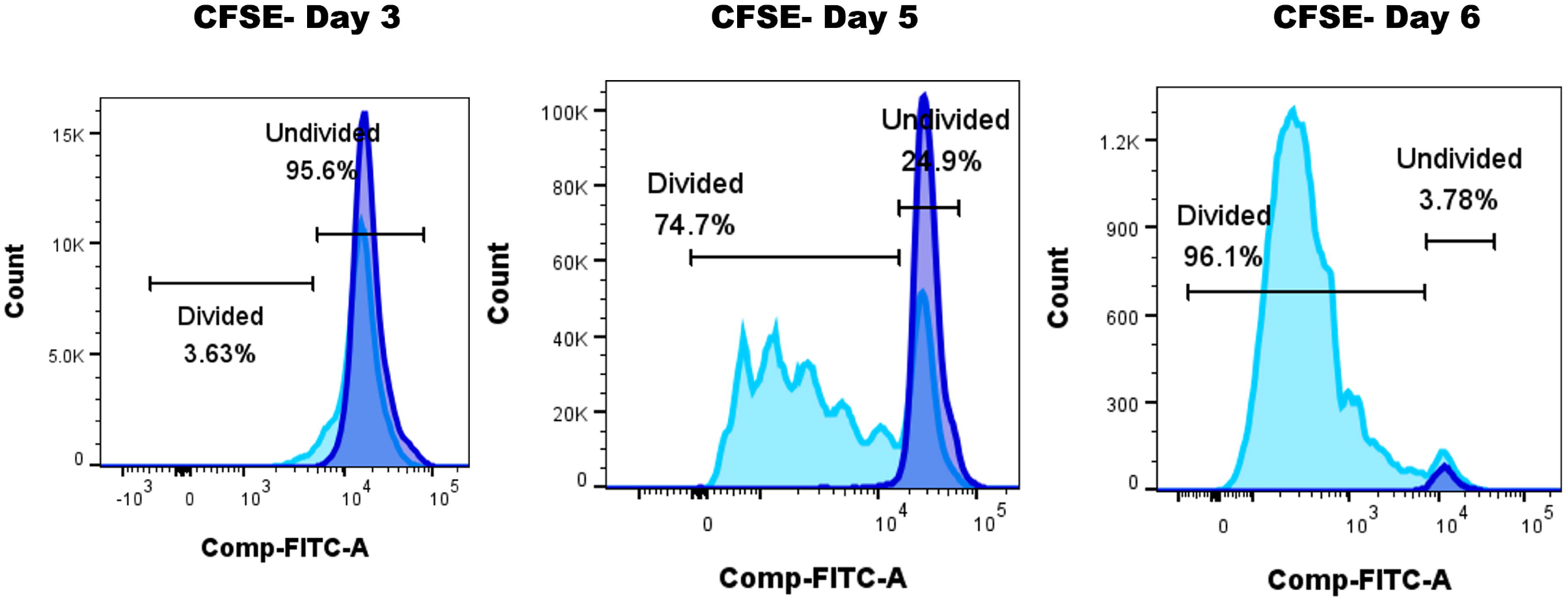
Optimisation of stimulation duration for maximal lymphocyte proliferation using CFSE assay.

### Ki-67 Staining and Time Point Determination

Titration of the Ki-67 antibody demonstrated that 2.5□μL per test provided optimal staining, characterized by low background fluorescence and clear separation between Ki-67□ and Ki-67□ populations. To determine the ideal duration of stimulation for capturing peak Ki-67 expression, 1□×□10□ PBMCs were stimulated with PHA and harvested at multiple time points: 24□h, 69□h, 72□h, 75□h, 78□h, 81□h, 96□h, and 120□h. Ki-67 expression gradually increased beginning at 24□h, plateaued between 69 and 96□hours with minor fluctuations, and declined thereafter likely due to activation-induced cell death or cellular exhaustion. Based on this kinetic profile, an incubation period of 72–75□hours was selected for subsequent Ki-67 assays, offering an optimal balance between assay sensitivity and turnaround time for assessing immune function in patient samples (Figure 3).

**Figure 3:**
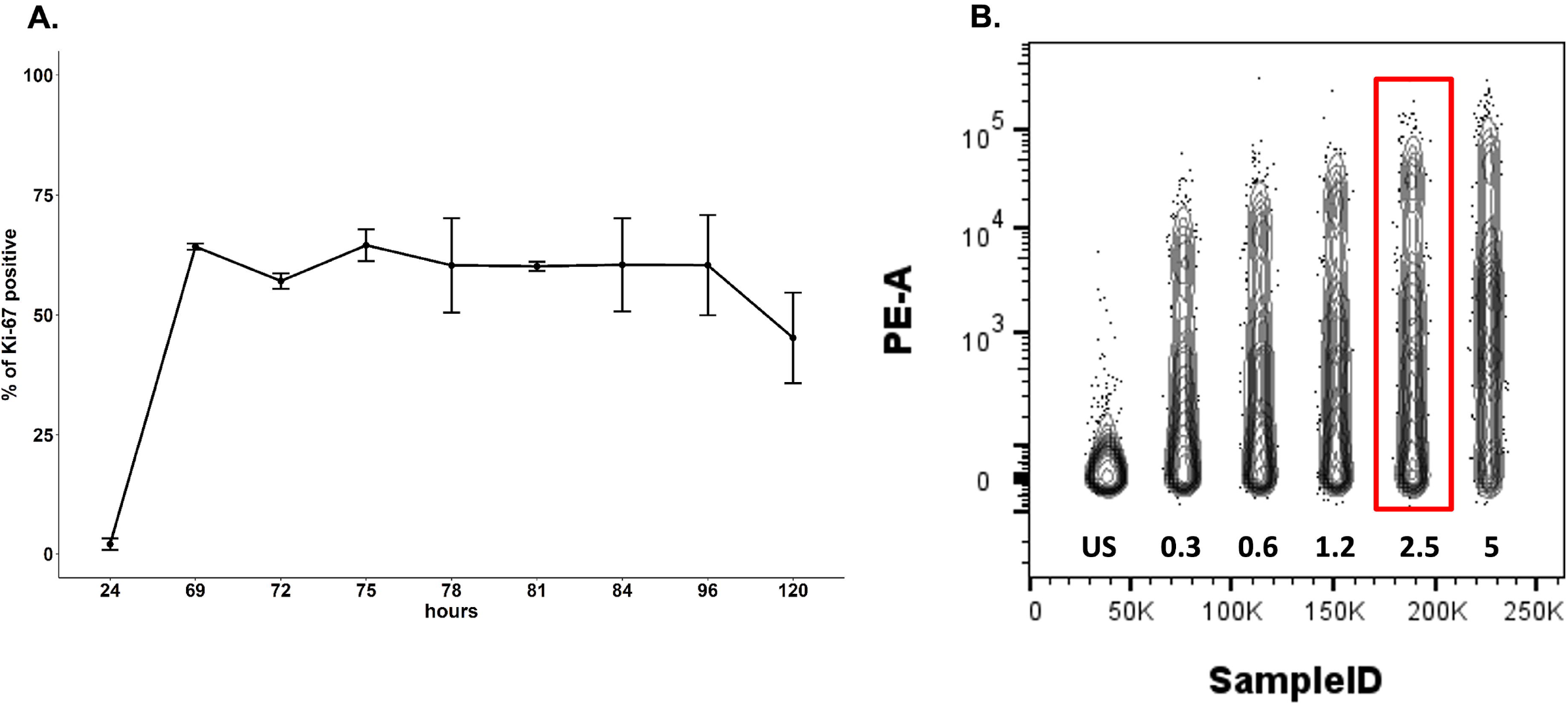
Optimization of Ki-67 staining for time kinetics and antibody titration. A) Time-course analysis of Ki-67 expression in stimulated lymphocytes at multiple time points post-stimulation. Each time point was assayed in duplicate across two independent experiments. Data are represented as mean ± standard deviation, reflecting the percentage of Ki-67□ proliferating cells over time. B) Concatenated image demonstrating the titration of Ki-67 antibody at varying volumes to determine the optimal concentration for maximal signal-to-noise ratio. The percentage of Ki-67□ cells is plotted to identify the volume yielding the most robust and specific staining without oversaturation or background signal.

### Effect of Cell Concentration on lymphocyte proliferation capability

To evaluate the effect of cell seeding density on lymphocyte proliferation assays, PBMCs were plated at four concentrations: 5□×□10□, 2.5□×□10□, 1□×□10□, and 0.1□×□10□□cells/mL. Among these, 1□×□10□□cells/mL yielded optimal results, with robust proliferation and clear resolution of CFSE peaks, while maintaining cell viability. In contrast, lower cell densities resulted in weak proliferative responses, and higher densities led to increased cell death and overlapping CFSE peaks, reducing the clarity of division profiles (Figure 4). Importantly, this seeding density also allowed for efficient use of limited sample volumes with lower PBMC cell counts, making it well-suited for assays in the paediatric population.

**Figure 4:**
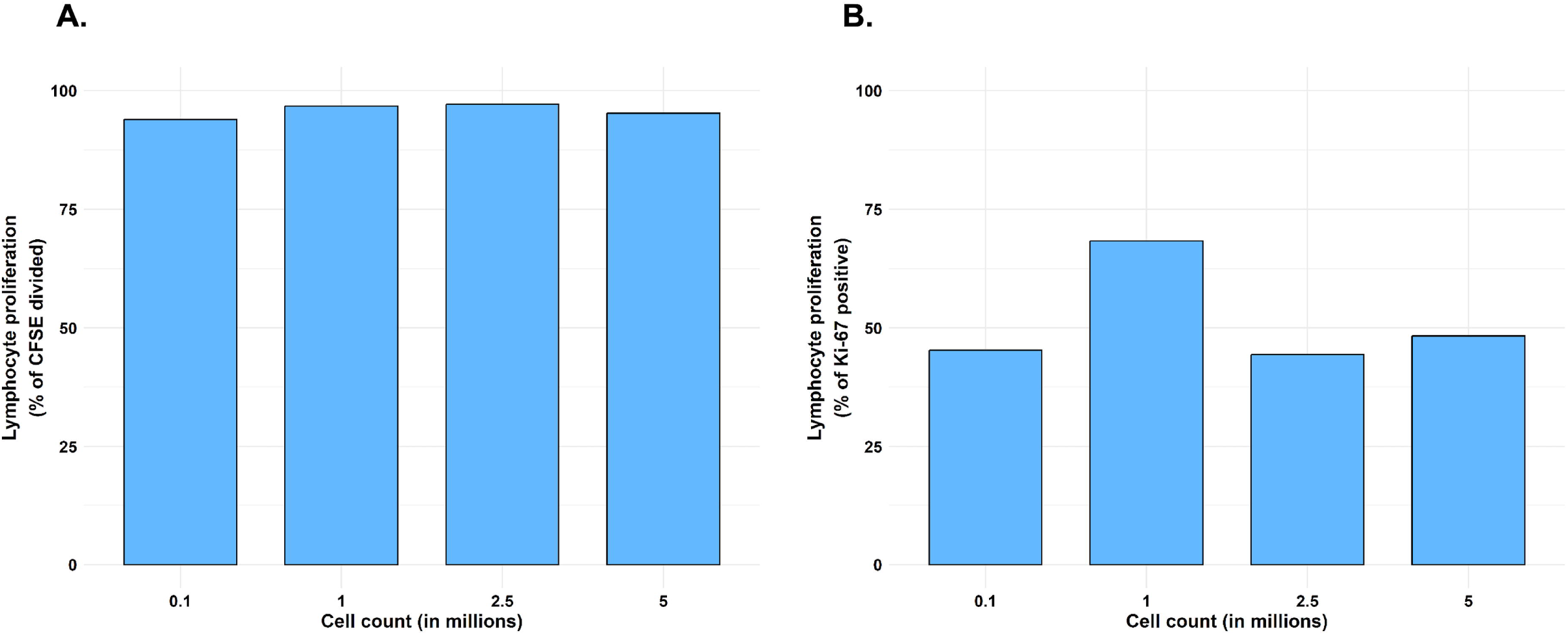
Effect of cell concentration on lymphocyte proliferation. A) Percentage of divided T cells as measured by CFSE dilution after 5 days of PHA stimulation across different cell concentrations (5□×□10□, 2.5□×□10□, 1□×□10□, and 0.1□×□10□□cells/mL). B) Percentage of Ki-67□ CD3□ T cells following PHA stimulation across the same range of cell concentrations.

### Participant Characteristics and Group Stratification

PBMCs were obtained from 20 pediatric participants (13 females). Participants were stratified into four clinical categories: well children (n = 3), nephrotic syndrome on corticosteroids (n = 12), off corticosteroids (n = 3), and children immediately post autologous stem cell transplant, presumed to be severely immunosuppressed (n = 2). The median age was 9 years (range 3-15 years). Demographic and treatment histories were recorded to explore potential differences in proliferative capacity (Table 1).

### Proliferative Responses Measured by CFSE and Ki-67

Both CFSE dilution and Ki-67 expression were used to quantify T cell proliferation for all the samples collected from the paediatric population. Representative histograms showed clear CFSE dilution in CD3+ T cells after 6 days of PHA stimulation demonstrating the cell proliferation (Figure 5) and the unstimulated cells showed negligible proliferation. Similarly, intracellular Ki-67 staining demonstrated a marked increase in CD3+ Ki-67+ cells at 72 hours post-stimulation, confirming active proliferation. Figure 5 shows the CFSE and Ki-67 based cell proliferation among the healthy and the child who was on steroid and did not show a good proliferation. The gating strategy for both the assays are shown in supplementary figure 1.

**Figure 5:**
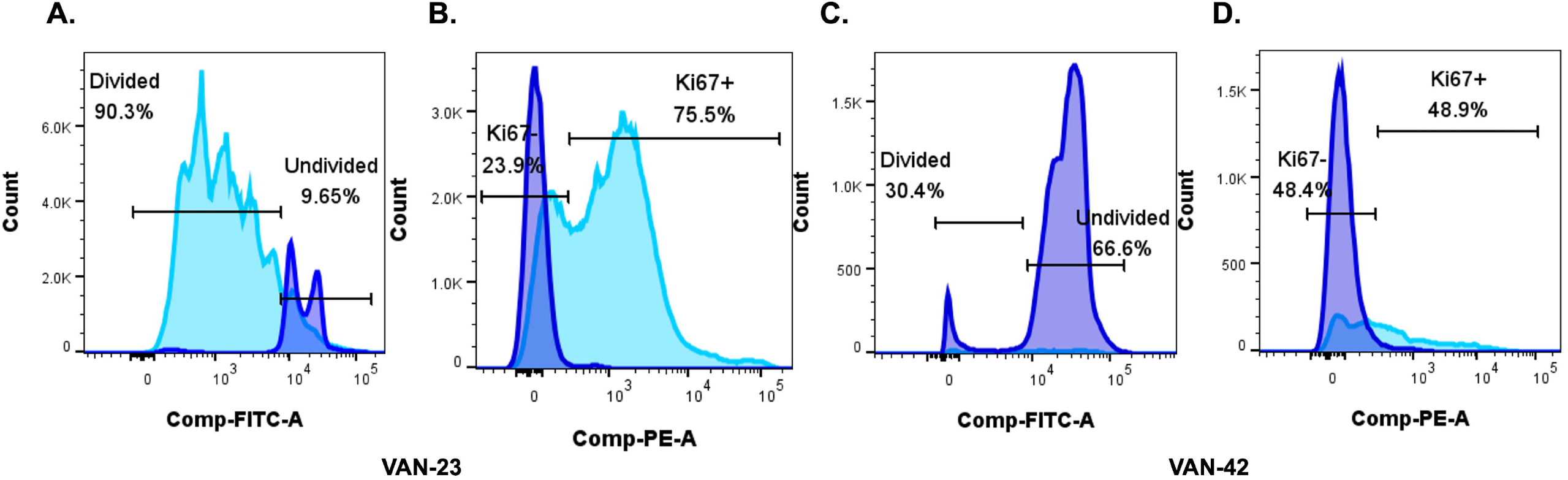
Lymphocyte proliferation assessed by CFSE dilution and Ki-67 expression in a healthy child and a child with nephrotic syndrome on corticosteroids. Lymphocyte proliferation was evaluated using CFSE and Ki-67–based assays following PHA stimulation. (A, B) CFSE dilution (A) and intracellular Ki-67 expression (B) in PBMCs from a healthy child show robust lymphocyte proliferation. (C, D) CFSE dilution (C) and Ki-67 expression (D) in PBMCs from a child with nephrotic syndrome on corticosteroid therapy reveal reduced proliferative capacity.

Quantitative analysis across multiple stimulated samples demonstrated a median percentage of proliferating CD3+ T cells ranging from 81.1% to 86.4% as measured by CFSE dilution. Ki-67 expression among CD3+ T cells ranged from 53.3% to 65.9% after background subtraction from unstimulated controls. A moderate positive correlation was observed between CFSE and Ki-67 based assays (Spearman’s r = 0.57, p = 0.01), indicating that both methods provide complementary assessments of T cell proliferation.

### Comparison across Clinical Groups

While the CFSE-based assay did not reveal marked differences in proliferative capacity across groups, the Ki-67 assay unexpectedly showed a higher percentage of proliferating CD3□ T cells in children receiving steroid therapy [median (IQR): 65.9% (61.2–69.5)] compared to healthy controls and children with nephrotic syndrome who were off steroids (Figure 6B). Although formal statistical comparisons were not performed due to the limited sample size, this trend suggests preserved or potentially enhanced T cell activation in the context of steroid treatment.

**Figure 6:**
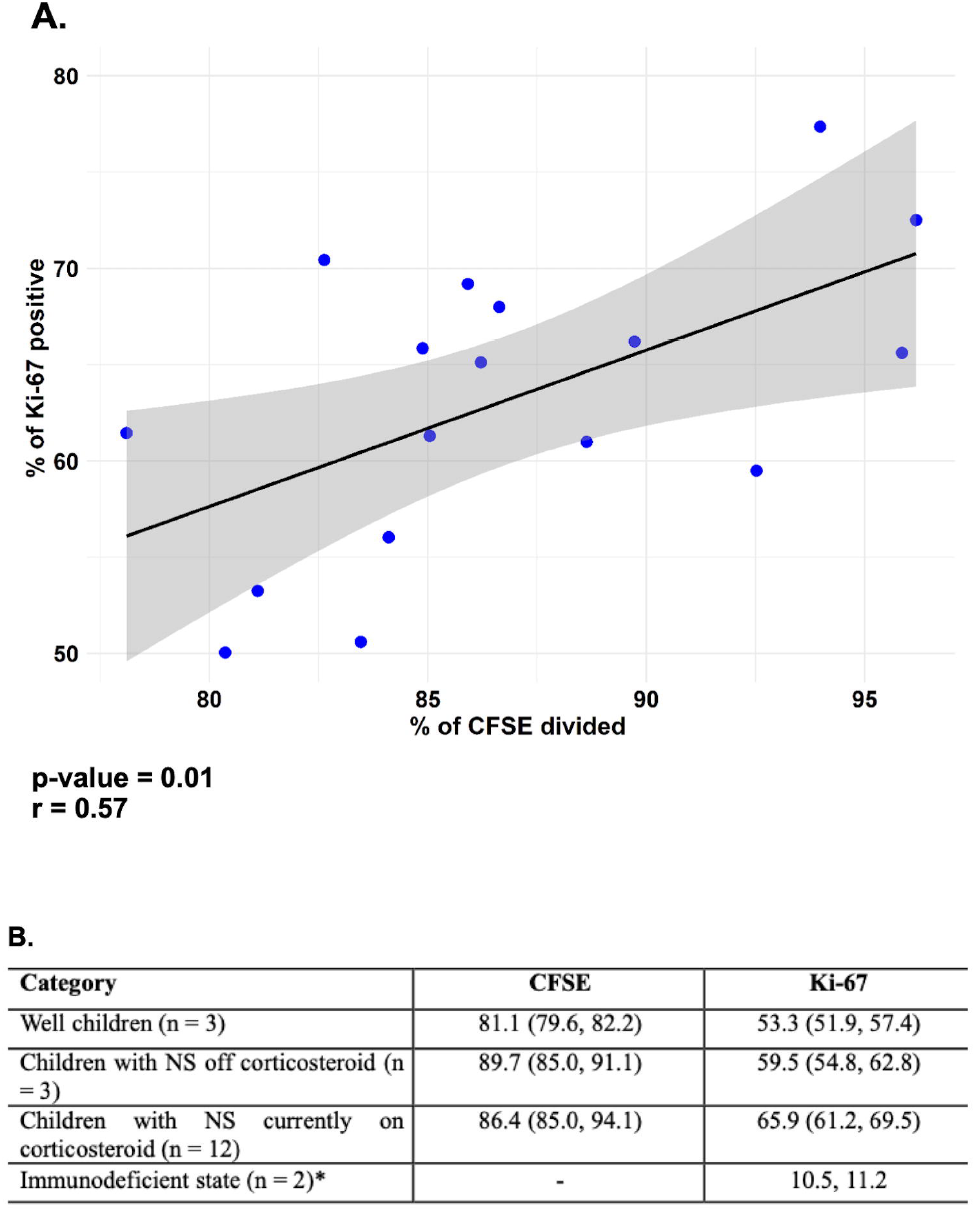
A. Correlation plot between Ki-67 and CFSE based lymphocyte proliferation comparison of CFSE and Ki-67–based lymphocyte proliferation assays in pediatric patients. B. Table summarizing the percentage of proliferating CD3+ T cells measured by CFSE dilution and Ki-67 staining across various pediatric patient groups. Data are presented as median with interquartile range *\*CFSE were not performed for the immunodeficient children due to the unavailability of adequate PBMC*. *NS-nephrotic syndrome, CFSE-carboxyfluorescein succinimidyl ester*

Overall, both CFSE and Ki-67 assays provided complementary and robust measures of T cell proliferation in pediatric PBMCs. These findings are particularly relevant for children with nephrotic syndrome receiving immunosuppressive therapy, where immune functionality appears to remain intact. The optimized, standardized protocols developed in this study offer practical and reliable alternatives to traditional thymidine-based assays, enabling broader application in low-resource research and clinical settings. Based on previous studies, a cut-off of >50% proliferation after background subtraction was used to define a positive proliferative response in both CFSE and Ki-67–based assays [9].

## Discussion

This pilot study optimized and validated two widely used lymphocyte proliferation assays, CFSE dye dilution and intracellular Ki-67 expression in cryopreserved PBMCs from a pediatric cohort with and without nephrotic syndrome in South India. A CFSE concentration of 2.5 μM was found to provide optimal fluorescence intensity while maintaining cell viability. We observed relatively preserved lymphocyte proliferation in children with nephrotic syndrome on corticosteroids when compared to those not on corticosteroids. Given that both PHA stimulation and CFSE labelling can be cytotoxic to PBMCs, it is essential to balance dye concentration to preserve cell integrity while enabling accurate proliferation tracking. Similar findings have been reported in earlier studies, which recommend CFSE concentrations between 1–5 μM for optimal resolution with minimal toxicity [4,10].

A cell density of 1 × 10□ cells/mL was chosen as optimal for labelling and stimulation. This is particularly advantageous in pediatric studies, where minimal blood volume is critical and repeated sampling is often limited. The CFSE-based assay showed a robust proliferation response at day 6 post-stimulation, consistent with the expected timeline for peak T cell expansion [11]. Unlike some previous studies that use shorter stimulation durations [12], we allowed sufficient time for stimulation to ensure that CFSE dilution reached levels comparable to unstained controls. This was critical to fully capture the proliferative capacity of lymphocytes, particularly in nephrotic syndrome children on corticosteroids, where immune function may be variably suppressed. Accurately assessing immunoproliferation in such a context requires complete division cycles to be represented.

Ki-67, a nuclear antigen expressed during all active phases of the cell cycle (G1, S, G2, and M), offered practical advantages for high-throughput analysis. Its expression remained relatively stable between 69 to 96 hours post-stimulation, allowing greater flexibility in sample processing and batching. However, for operational feasibility and timely distribution of preliminary patient reports, we intend to perform Ki-67–based stimulation assays within 72–75 hours post-stimulation, a window that still captures peak expression reliably based on our findings. Compared to CFSE, Ki-67 assays require fewer pre-labelling steps and are compatible with cryopreserved cells, making them especially suitable for large-scale or multicenter studies [13].

A moderate but statistically significant correlation (Spearman’s r = 0.57, p = 0.01) was observed between CFSE dilution and Ki-67 expression, reflecting their shared but non-identical capacity to capture cell proliferation. CFSE detects actual cell divisions through dye dilution, while Ki-67 identifies cells in the cell cycle regardless of how many divisions have occurred. Studies have shown that while each method independently provides useful information, their combined use improves interpretability and reliability of proliferation data, particularly in heterogeneous samples or in clinical cohorts with variable immune responsiveness [9,13].

Importantly, performing both assays within the same experimental setup enhances confidence in results through internal validation. Discrepancies between the two markers can offer additional biological insights, for example, cases where Ki-67 expression is high but CFSE dilution is minimal, may reflect early activation without progression through multiple divisions. Using both assays also improves assay robustness in settings where one marker may be technically compromised (e.g., poor CFSE staining or suboptimal Ki-67 fixation). While slightly more labor-intensive, combining both readouts in a single stimulation setup is cost-effective compared to running independent assays on different days and conserves precious PBMC samples, especially from pediatric participants.

In summary, we demonstrate that the use of both CFSE and Ki-67 assays is feasible, complementary, and robust for assessing T cell proliferation in cryopreserved PBMCs. These findings will be leveraged in upcoming studies evaluating live attenuated vaccine responses in immunocompromised children, such as those with nephrotic syndrome on corticosteroids, where accurate assessment of immune function is essential. Our results provide a reproducible, safe, and scalable protocol suitable for pediatric immunological studies in resource-limited settings. The assays are applicable for disease monitoring, pre-evaluation screening before administering live vaccines, and immunomodulatory studies, particularly in children undergoing immunosuppressive therapy.

Before broader implementation, it is essential to test and confirm the robustness and reproducibility of this protocol in diverse cohorts, including children with varying immune statuses and clinical backgrounds. Future work will validate these findings in larger, diverse cohorts and evaluate antigen-specific proliferation responses following vaccination.

## Data Availability

All data produced in the present study are available upon reasonable request to the authors

## Competing Interests

The authors have no relevant financial or non-financial interests to disclose.

### Ethics approval

Informed consent was obtained from guardians of all participating children, and the study was approved by the institutional ethics committee (IRB Min. No. 2411130)

## Statements and Declarations

### Author Contributions

G.M and I.A conceived the trial and contributed to the original protocol. G.M, contributed to the project administration and data entry. C.S, P.S, A.K performed the CFSE and Ki-67 assays. R.M provided the scientific guidance for the assay standardization. A.E performed the statistical analysis. E.O acquired the samples by flow cytometry, L.L.J provided samples from immunodeficient patients. S.K.L provided critical inputs for the assays. R. M. drafted the initial report. G.M and R.M finalized the report. All authors reviewed and approved the final report.

### Data, Material and/or Code availability

Data sets generated during the current study are available from the corresponding author on reasonable request.

