## Supplemental figures for "Standardization of CFSE and Ki-67 based lymphocyte proliferation assays in the absence of thymidine-based methods among children with nephrotic syndrome in south India: A pilot study": Supplementary figures and tables.docx

**Supplementary figure 1**: **Gating strategy for assessing lymphocyte proliferation following PHA stimulation.** (A) Gating strategy for detecting Ki-67 expression in proliferating T cells. (B) Gating strategy for evaluating CFSE dilution–based proliferation. In both panels, gating was performed sequentially on lymphocytes (based on FSC/SSC), live cells (FVS700⁻), and CD3⁺ T cells, followed by assessment of proliferation using either intracellular Ki-67 expression or CFSE dye dilution.

**A.**

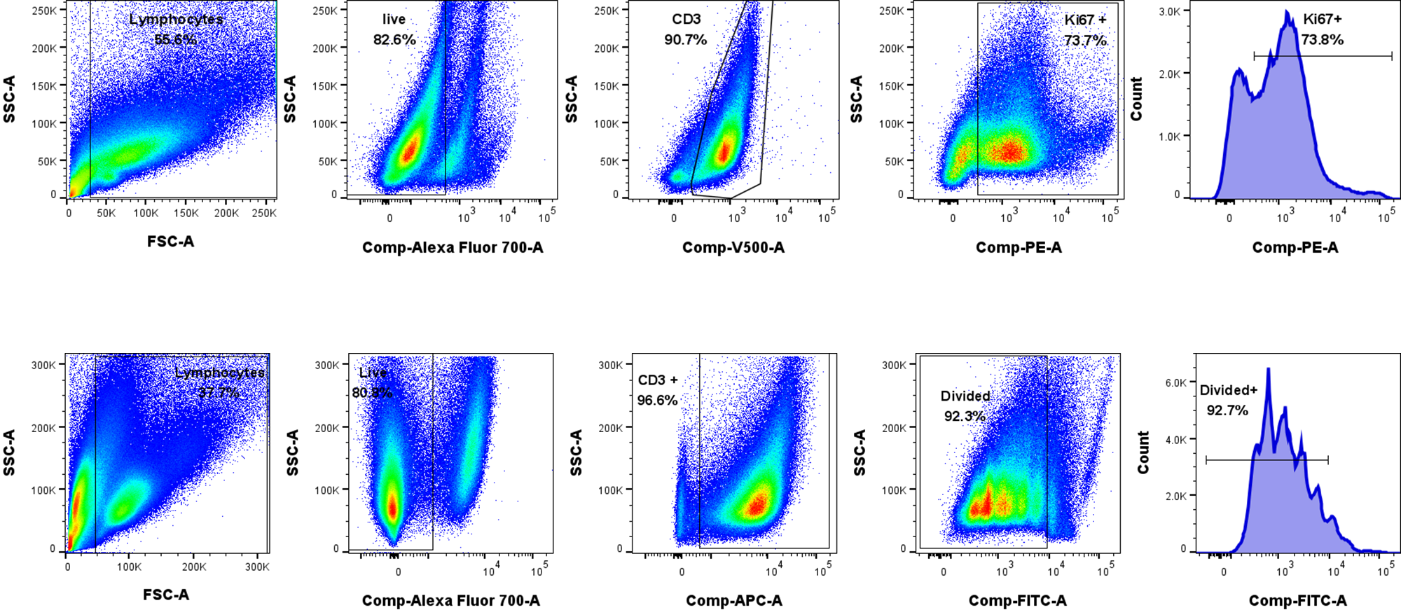

**B.**

**Supplementary figure 2: Inter-assay variation assessing the repeatability of CFSE-based lymphocyte proliferation assay.** Lymphocyte proliferation following PHA stimulation was evaluated using CFSE dilution in two independent experiments (Trial 1 and Trial 2) conducted on different days using the same cryopreserved PBMC sample. Each trial included parallel testing of 1 × 10⁶ cells with two CFSE concentrations (2.5 µM and 5 µM). Consistent CFSE dilution patterns were observed across both trials and conditions, demonstrating good assay reproducibility and robustness.

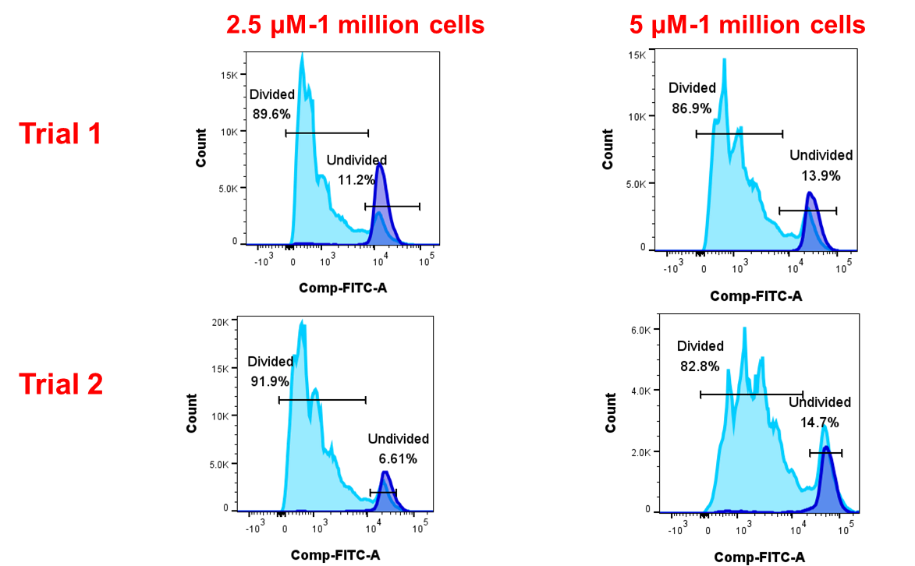
